# Changing case fatality risk for COVID-19 over time in selected European countries

**DOI:** 10.1101/2020.11.26.20239327

**Authors:** Heiko Becher, Katharina Olszewski, Sarah Wiegel, Olaf Müller

## Abstract

**Objectives:** To illustrate the development of the case fatality risk (CFR) for COVID-19 over time using different assumptions for calculating the CFR.

**Design:** Observational study.

**Setting:** 

Selected European countries, 28 January to October 29 2020.

**Participants:** Laboratory-confirmed COVID-19 cases and deaths due to COVID-19

**Main outcome measure:** case fatality risk (CFR)

**Results:** We show that the CFR has considerably decreased over time. This seems to be driven not only by increased testing but also by a reduced CFR among cases older than 60 years. Our data also confirm a significantly higher fatality risk for men than for women. The decline in the CFR is even more pronounced when only cases and deaths occurring in a specified time window are considered. This alternative estimation method has the advantage that early data where the bias due to the incomplete ascertainment of cases was arguably largest do not affect CFR estimates later on. We find similar results for other European countries.

**Conclusion:** CFR estimates vary considerably depending on the underlying assumptions concerning their calculation. Reliable CFR estimates should not be based on cumulative numbers from the beginning of the pandemic but rather be based on more recent data only.

## Introduction

The outbreak of coronavirus disease 2019 (COVID-19), which is caused by the severe acute respiratory syndrome corona virus 2 (SARS-CoV-2), has started in China by the end of 2019 (1, 2). It has rapidly developed into a serious pandemic, and by November 3, a total of 46.871.264 cases were reported globally, with a total of 1.206.187 deaths (3, 4). Until today, most cases and deaths occurred in the USA, India, Europe and Latin America, with a second wave of the epidemic now rapidly emerging in all countries of the northern hemisphere (3, 4).

A central parameter to assess the burden resulting from SARS-CoV-2/COVID-19 is the case fatality risk (CFR). Many publications report the crude CFR, calculated as the cumulative number of deaths divided by the cumulative number of cases. Yet, estimation of the crude CFR may be affected by several biases. More sophisticated methods are available but their application is often hampered by a lack of detailed data (5, 6).

In an earlier paper, we have shown that the different age distribution of cases in various European countries can explain a large proportion of observed differences in CFRs (7). Furthermore, dividing deaths by the number of cases at a certain point in time neglects the delay between infection and reported death. Thus, the denominator, i.e. the number of cases, should include a certain time lag so that only cases that allow for a sufficiently long follow-up time for a potential death to occur and be recorded are included in the analysis. According to the World Health Organization (WHO), time between symptom onset and death ranged from 2 to 8 weeks (8). Limited testing capacity, in particular at the beginning of the pandemic, made it difficult to estimate the denominator of the CFR. A number of studies reported varying numbers of asymptotic Covid-19 infections: the estimates range from 18 % among the passengers of the cruise ship *Diamond Princess* (9) to 45 % in a two-point prevalence survey in the Italian city of Vo (10). Asymptotic or mild cases are less likely to be reported to the surveillance system. Therefore, the CFR estimate will be higher among ascertained cases than among the entire population of cases. Finally, the overwhelmed health system in Italy in March 2020, for example, resulted in a very high CFR due to the limited capacity of intensive care facilities in hospitals (11). While there is sufficient knowledge that the CFR of SARS-CoV-2/COVID-19 is strongly associated with old age and chronic diseases, there is less knowledge on other related factors (5, 2).

The incomplete ascertainment of cases was particularly relevant during the early phase of the pandemic when testing capacity was low (2). If these underreported cases remain as cumulative numbers in later estimates, this bias somewhat decreases, but may still have a large impact on the CFR estimate. To avoid this bias, we present an alternative estimation method in this paper. We compare the crude and age- and sex-adjusted CFR estimates both using cumulative case and death numbers as well as cases and deaths occurring in a more recent time window. We also consider estimates by sex and age group.

In the light of our findings we discuss three further aspects which may have an effect on the CFR estimates: (a) the increase of the detection rate of the cases due to increasing testing capacities (12), (b) the improved treatment options for Covid-19 (13), and (c) mutations of the virus that may have led to changing lethality (14, 15).

## Data and Methods

Data on daily age-and sex-specific case and fatality numbers in Germany until October 29 were obtained from the Robert-Koch-Institute (RKI) (16). The RKI disaggregated case and fatality numbers by sex and age, presented in six age groups j=1, …,6 as 0-4 years, 5-14 years, 15-34 years, 35-59 years, 60-79 years, and 80 years and above. International data on daily case and fatality numbers until October 29 were obtained from the European Centre for Disease Prevention and Control (17). These data are not separated by age group and sex. We restrict our analysis on European countries which accumulated more than 5000 deaths until October 28, 2020. These are Belgium, Germany, France, Italy, Netherlands, Romania, Russia, Spain, Sweden, Ukraine, UK.

We estimated the CFR for COVID-19 for the period April 1 until October 29 by using the following four methods:

i. cumulative crude CFR (cumulative number of deaths up to “date” divided by cumulative number of cases up to “date minus k days”)

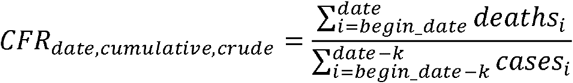
ii. cumulative age-standardized CFR (Germany only) (as (i), however age-standardized)

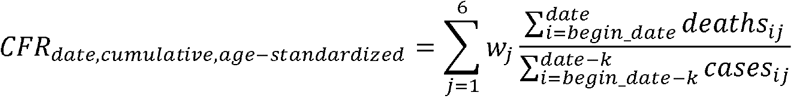
iii. 60-day crude CFR (60-day crude CFR (number of deaths in the last 60 days before “date” divided by number of cases in days “date-60-k” until “date-k”)

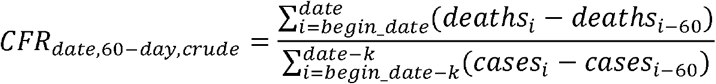
iv. 60-day age-standardized CFR (Germany only) (as (iii), however age-standardized)

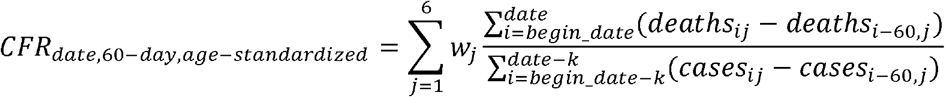

In addition, we calculated the CFR by age group and sex for Germany. The time delay between case reporting and death was initially assumed to be 7 days. A longer delay of 14 days was also considered for comparison. As “begin_date” we defined the first date of reporting a case which is January 28, 2020 for Germany. The weights *w*_*j*_ for age standardization were chosen according to the standard European population 2013 (18). All calculations were performed using the statistical software package SAS.

Patient and Public Involvement

~~~
No patient involved
~~~

## Results

Figure 1 shows the crude and age-standardized CFR for Germany from April to October 2020, both using cumulative numbers and considering only the last 60 days of any given date. All CFR estimates decrease over time, although their development over time varies considerably. After a peak during the first wave with a crude and age-standardized CFR of 8% and 7%, respectively, the cumulative age-standardized CFR (red dashed line) rapidly decreases to about 3% and nearly plateaus thereafter. The cumulative crude CFR (blue dashed line) decreases slower but converges to a similar CFR estimate (around 4%) at the end of the considered time period. The higher case-fatality risk among the older population drives the larger crude CFR estimate. Cumulating cases and death only over the last 60 days at each point of time results in a much sharper decrease of CFR estimates. The age-standardized CFR estimate at the end of the observation period using a 60-day window is 0.006 (0.6%), about five-fold lower than the age-standardized estimate using the cumulative numbers. Since the age-standardized CFR decreases over time, the apparently low death numbers despite the rising number of cases over the last months cannot be attributed only to the changing age-distribution of the cases.

**Figure 1.**
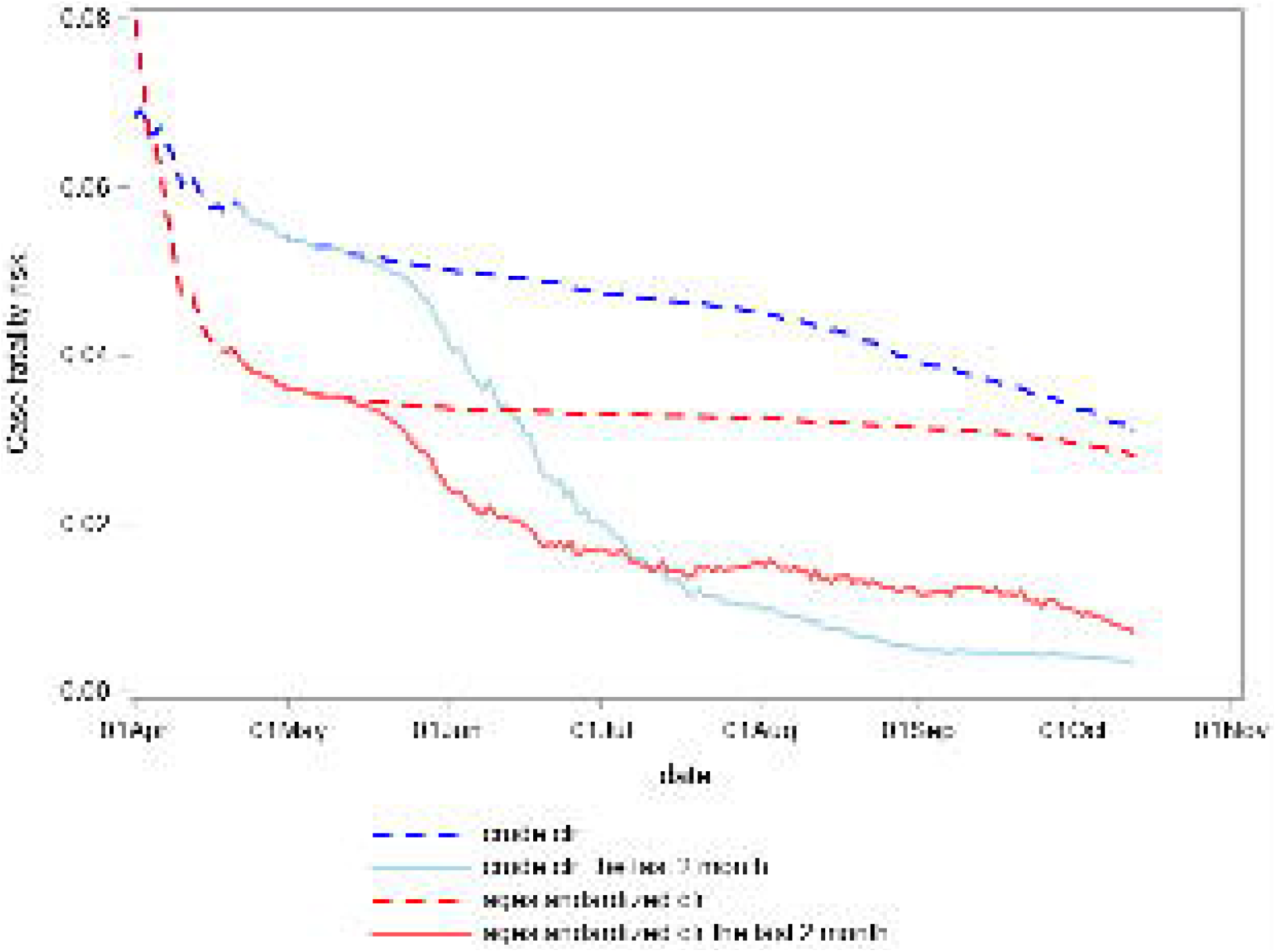
CFR estimates, 60-day time window, Germany

Figure 2 shows the age-standardized CFR estimates over time using a 60-day window, separately by sex. It confirms the observation from other studies that males have a higher death risk, with a factor of about two.

**Figure 2.**
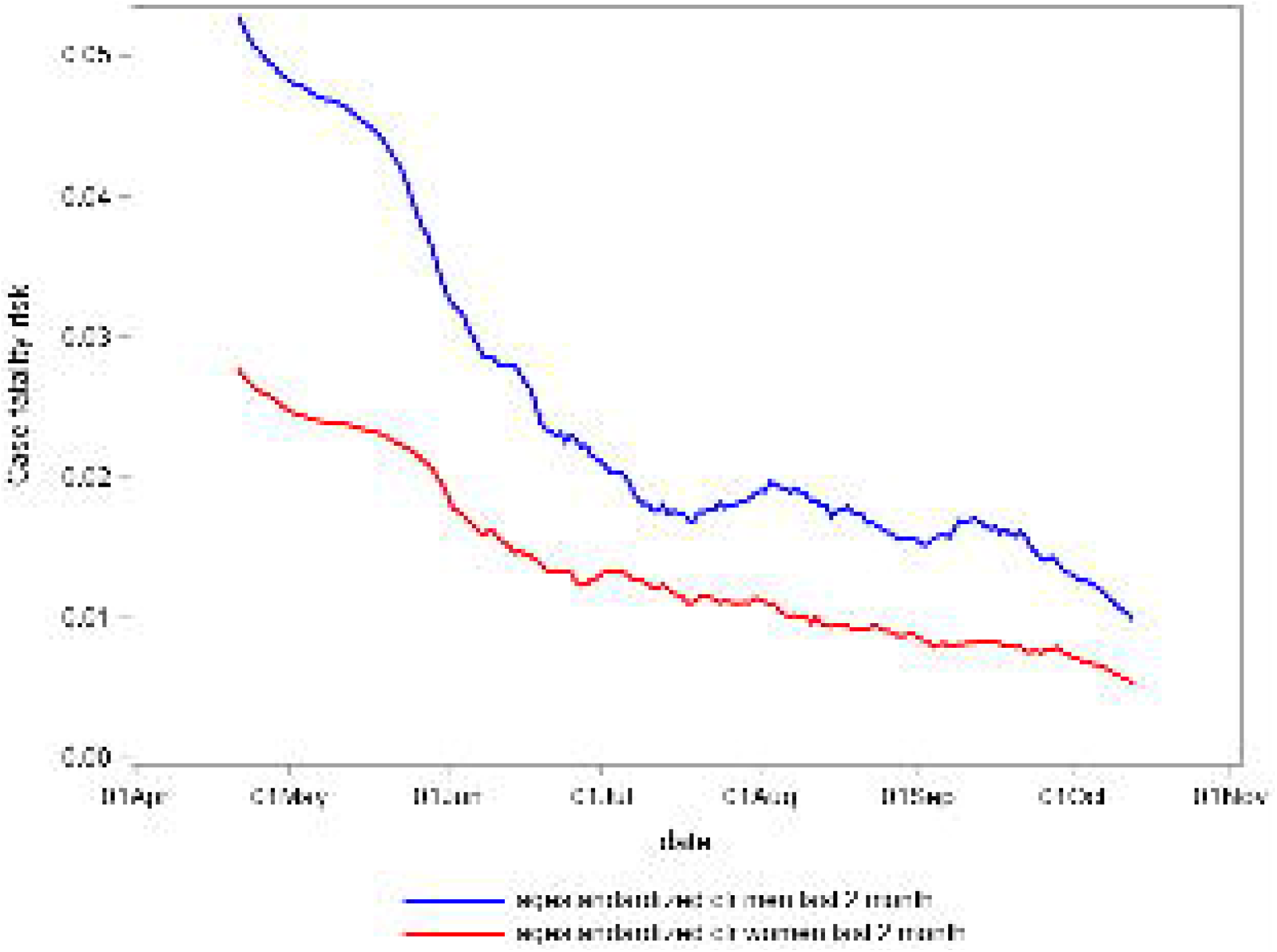
Age-adjusted CFR estimates by sex, 60-day time window, Germany

Figure 3 demonstrates the strong age-dependence of the CFR. The age-specific CFR estimates for the different age groups, 60-day window, are displayed. The age groups below 60 years (only the age group 35-59 is depicted here) have a constantly low CFR of 0,6% and below. For the age group 60-79 years, the CFR continuously decreased from May to 1.35% on October 13. The CFR of the age group 80 years and above fluctuates around 12% in the months of July to October after a steep decline in the months before.

**Figure 3.**
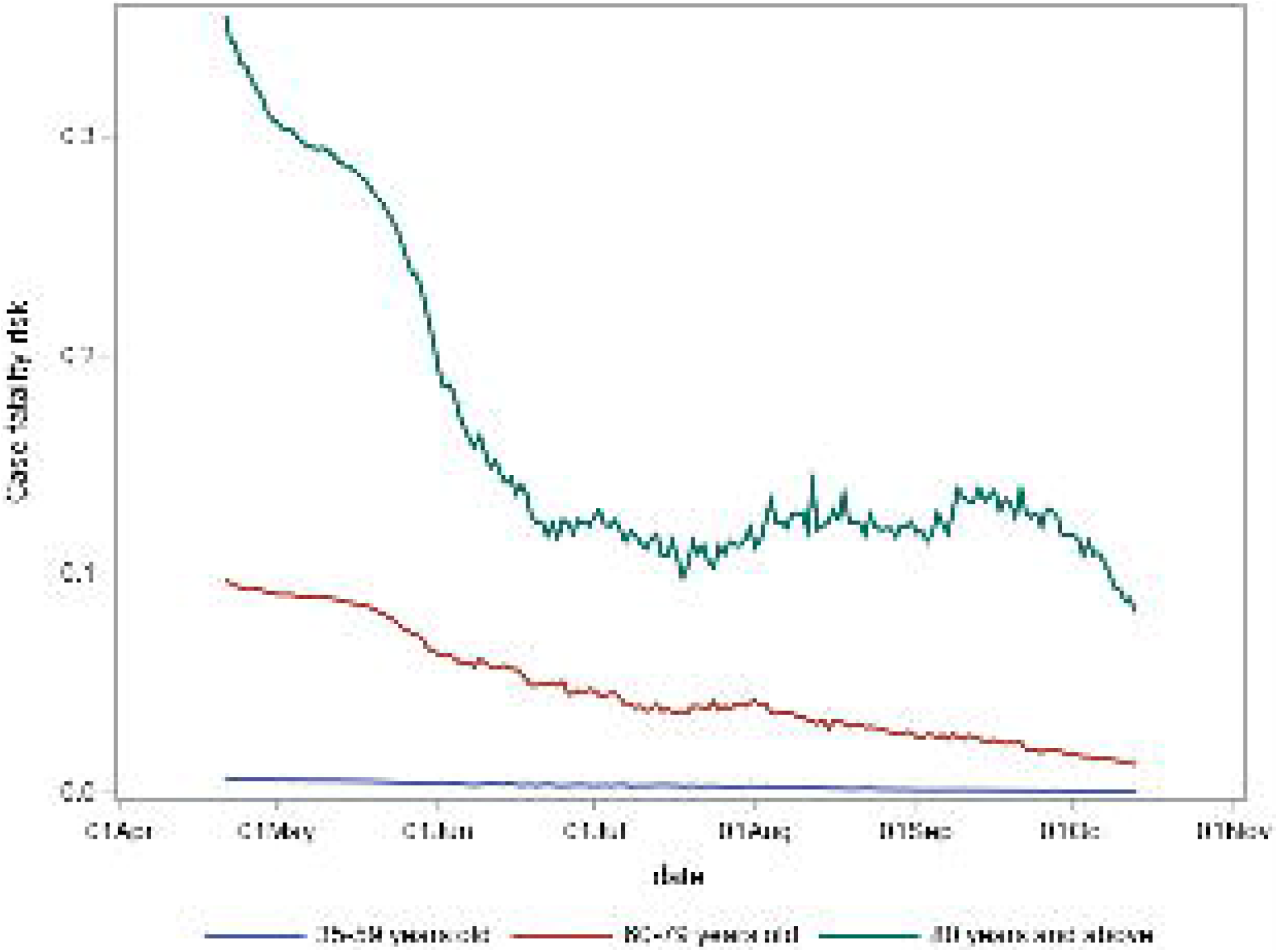
Age-specific CFR estimates, 60-day time window, Germany

Figures 4 and 5 show the crude risks for selected European countries which have more than 5000 cumulative deaths until October. Estimates in figure 4 are based on cumulative numbers, estimates in figure 5 are based on a 60-day window. There is some decrease in the estimates for all countries except Russia using the cumulative numbers; however, the decrease is relatively small. The time window data, on the other hand, show a different picture. While for the Eastern European countries (Russia, Romania, Ukraine) the differences are relatively low, we observe in figure 4 a strong decrease in the other countries (Belgium, France, Germany, Italy, Netherlands, Spain, Sweden, UK) with crude estimates close to each other and all below 1%.

**Figure 4.**
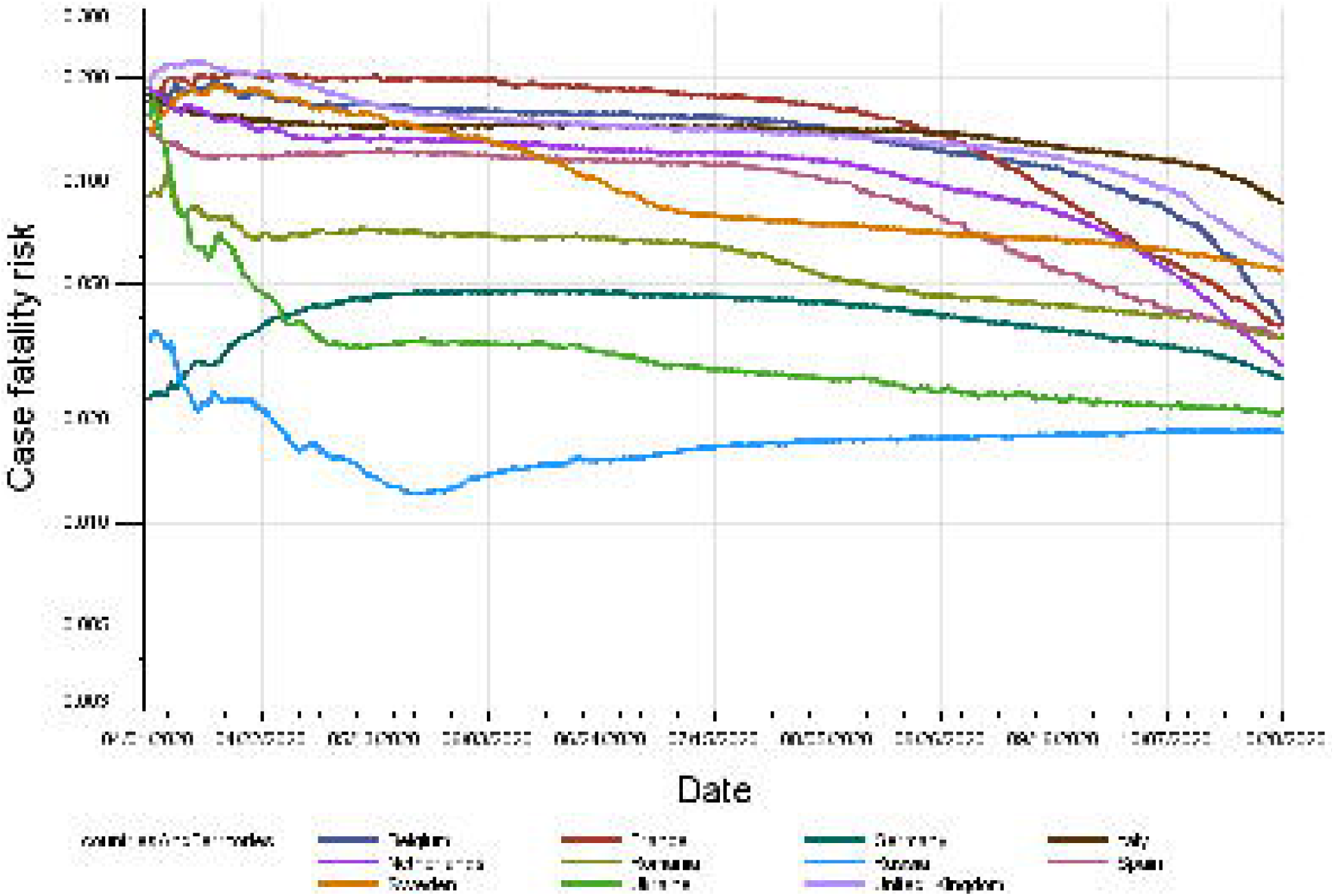
Crude CFR estimates, cumulative numbers, selected European countries

**Figure 5.**
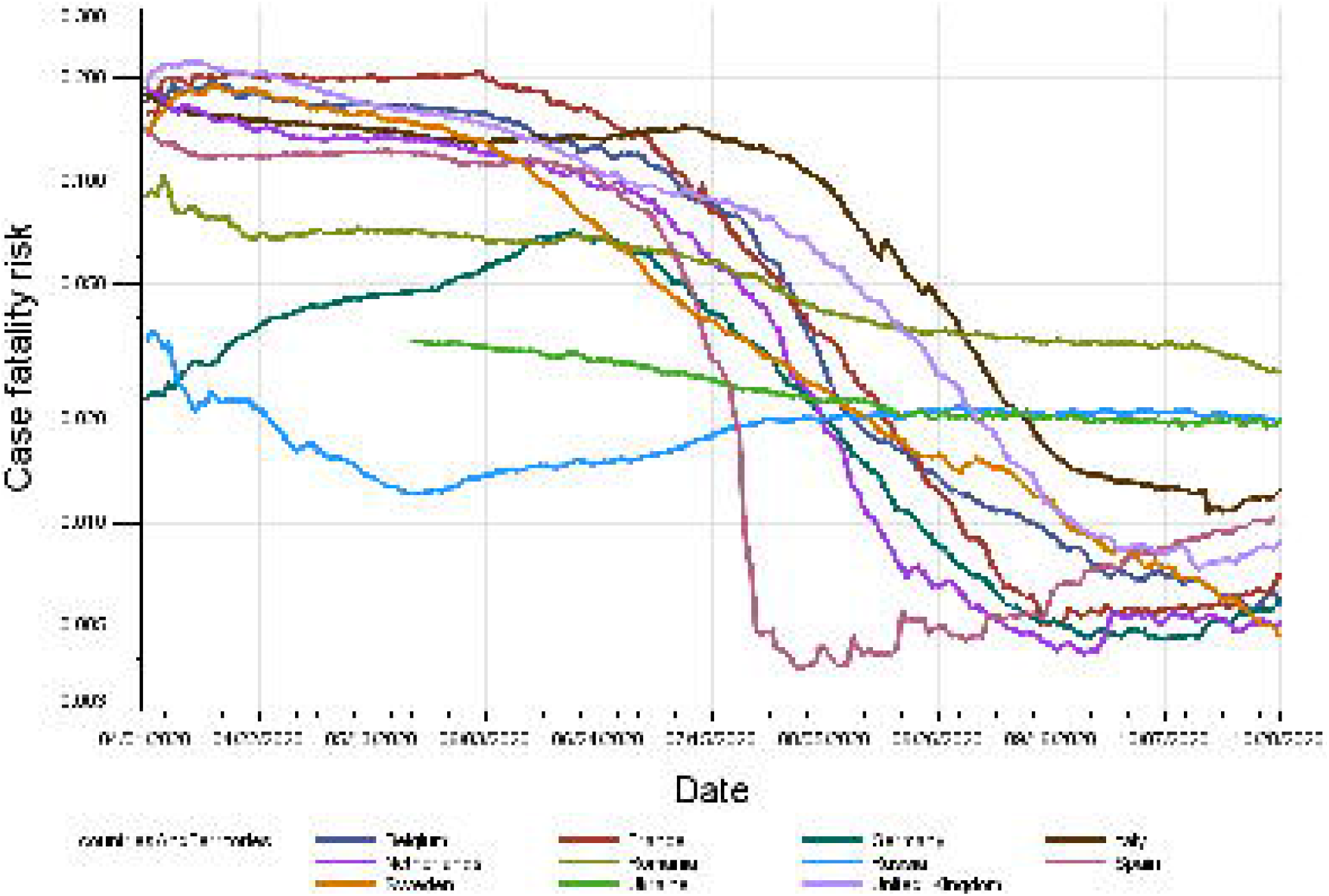
Crude CFR estimates, 60-day time window, selected European countries

Table 1 shows the estimates for the last date of our database, October 28, 2020. In this table, we also consider the effect on the CFR estimates when a longer period of 14 days from reporting a case to death is assumed instead of 7 days. In a period with daily rising numbers, such as in the second half of October, the age-standardized CFR estimate is notably higher when assuming 14 days rather than 7 days (1.27% vs. 0.79% on October 29) however still much lower than at the beginning of the pandemic. During the times when the numbers were relatively constant until, say, begin of October 2020 the differences in the CFR estimates with lag 14 or 7 days are smaller. On September 1, the values are 1.25% and 1.20% (ratio 1.04), and on October 1, the values are 1.19% and 1.00% (ratio 1.19).

**Table 1.**
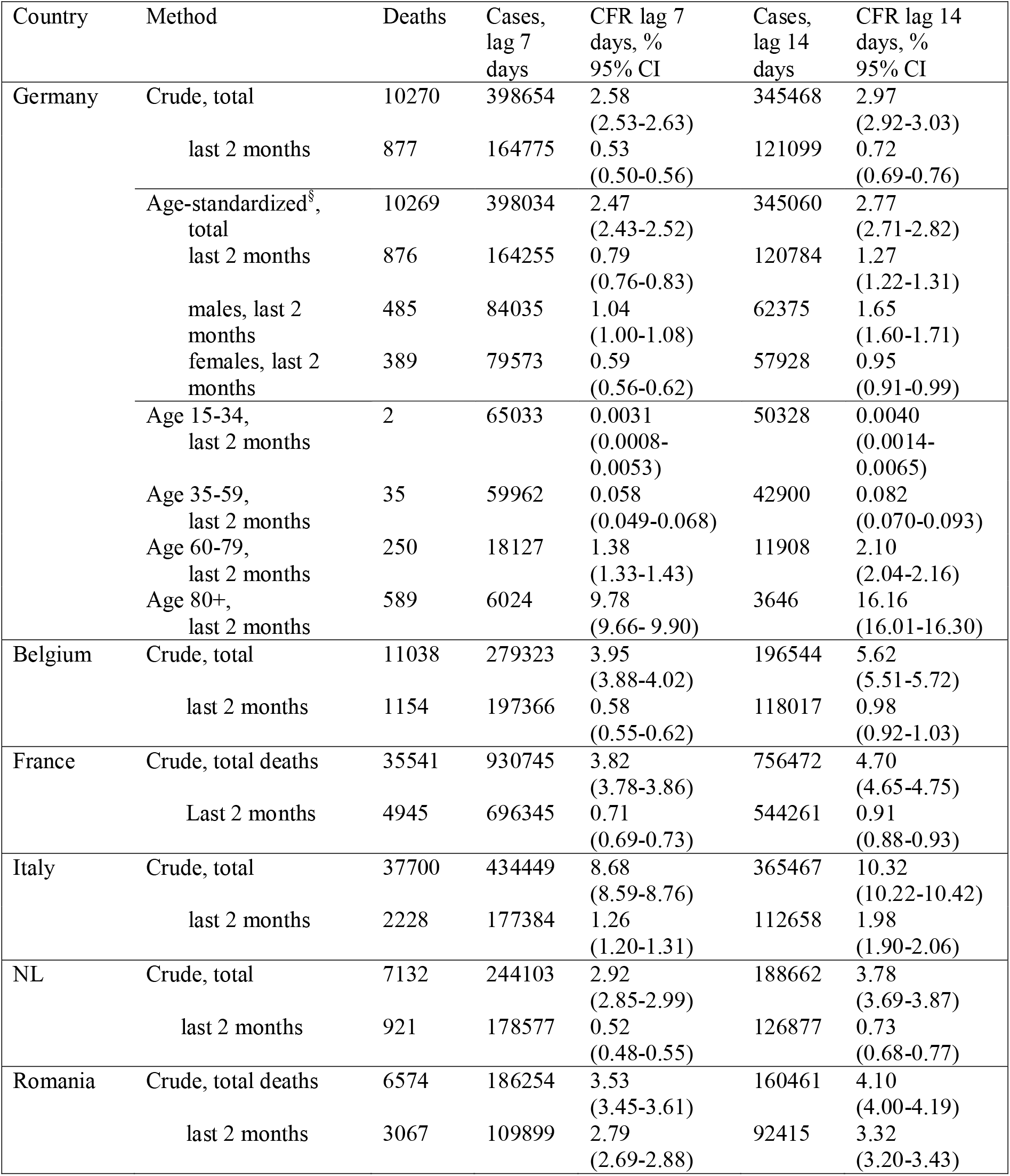
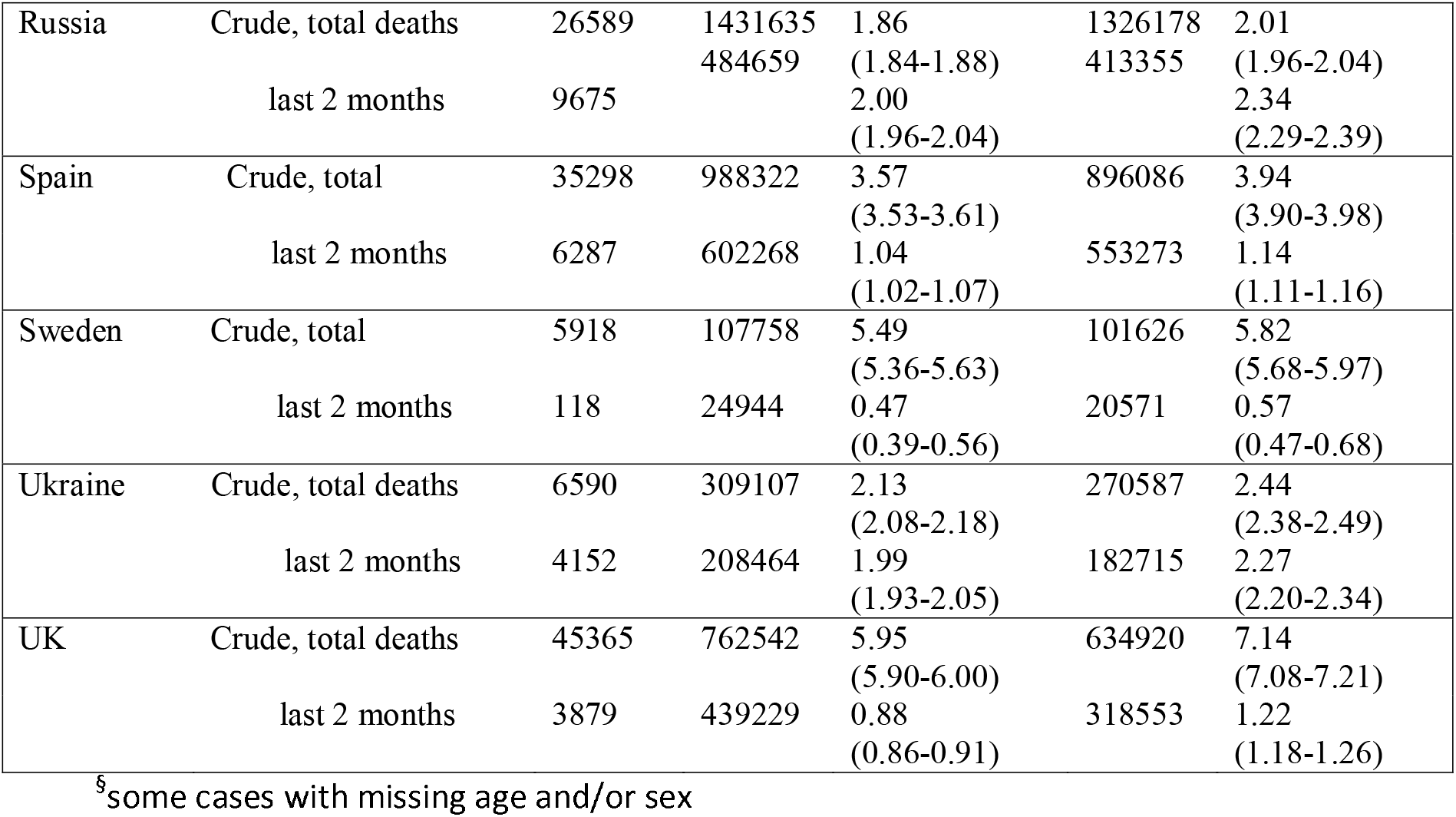
CFR estimates according to different estimation methods, Germany and selected European countries, October 29, 2020.

## Discussion

In Germany, we observed a reduction of the CFR over time which can only partly be attributed to the changing age distribution of the infected cases towards younger ages. We could also demonstrate that the use of cumulative cases and deaths from the beginning of the pandemic results in a strong overestimation of the CFR, both in Germany and in the other European countries analysed. Since there is no complete ascertainment of Covid-19 cases, our estimates based on the last 60 days may still be an overestimation of the true CFR.

A more complete ascertainment of SARS-CoV-2 infections and Covid-19 cases is only possible under special conditions of closed cohorts or in populations with additional comprehensive sero-surveys. A study on the closed cohort of the *Diamond Princess* cruise ship in Asia has provided an CFR of 0.99% (19, 9), while a representative population survey in Geneva/Switzerland demonstrated an IFR of 0.64% (20). The age-standardized CFR in the *Diamond Princess* population was calculated as 0.65%, which is close to the study from Switzerland and to our calculations from Germany (7). Preliminary results from a nationwide representative sero-survey in Spain showed an overall CFR between 1.1% and 1.4% in men and 0.58% to 0.77% in women (21). In this study, the CFR increased sharply after age 50, ranging between 11.6% and 16.4% in men ≥80 years and between 4.6% and 6.5% in women ≥80 years (21). Note that we do not distinguish between infection fatality and case fatality since asymptomatic cases, which occur in various testing settings of COVID-19, are also reported as cases.

Although our data support the many studies showing that the CFR attributed to SARS-CoV-2/Covid-19 is lower than initially suggested, these results cannot be taken as a message that COVID-19 is just a “light flu” as suggested by some (22, 5). An uncontrolled spread of the virus in European countries will certainly result in high numbers of hospitalized patients beyond healthcare capacity and an unacceptable large number of deaths (23).

It is challenging to disentangle the possible causes for the change of the CFR estimates over time. Possible causes for the observed decrease of the CFR are (i) an increase in the detection rate of cases, (ii) improved treatment options, or (iii) mutations of the virus. Testing capacity was limited at the beginning of the pandemic in all countries, and therefore symptomatic cases were more likely to be tested. Before July, about 300.000 to 400.000 tests per week were performed in Germany. In March/April, 6 to 9% of tests were positive, and this proportion decreased to under 1% in summer. From July onwards, test numbers increased and reached a plateau of about 1.1 million tests per week by now (24). Afterwards, the proportion of positive tests gradually increased and reached 5% in by the end of October. It may be appropriate to assume that the proportion of undetected cases is rather constant from, say, August. Furthermore, it is likely that case management has improved with increasing experience in the hospitals, and there is one medication (dexamethasone) which has been shown to significantly reduce the CFR in patients with severe Covid-19 disease (25). Finally, there is currently some but still rather limited evidence for changes in the genetics of the SARS-CoV-2 which may influence outcomes (14, 15, 26).

Our estimation methods included some assumptions as well as limitations. We aimed to keep the calculations simple using a fixed time from reporting to death. The distribution and its mean may have changed over time towards a shorter interval from infection to reporting. In a period with daily rising numbers, such as in the second half of October, the age-standardized CFR estimate is notably higher when assuming 14 days rather than 7 days. It is possible that a time of 7 days may be too short, which would yield to an underestimation of the CFR when the case numbers are quickly rising. Until mid-October the differences in the CFR estimates with lag 14 or 7 days are small, as shown in the results, and these estimates appear to be robust.

The time window of 2 months has been chosen arbitrarily. We selected an interval which is long enough to accumulate sufficient numbers for a stable estimation, and short enough to show changes over time clearly. We checked other intervals (one and three months) and found very similar results.

In conclusion, we have suggested an estimate of the CFR of COVID-19 which is based on more recent data only. We recommend not to calculate CRF based on cumulative numbers from the beginning of the pandemic. We showed a decrease of the CFR of COVID-19 over time.

## Data Availability

Data are freely available under
https://npgeo-corona-npgeo-de.hub.arcgis.com/datasets/d4580c810204019a7b8eb3e0b329dd6_0/data.
and
https://www.ecdc.europa.eu/en/publications-data/download-todays-data-geographic-distribution-covid-19-cases-worldwide.

## Article Summary

### Strengths and limitations of this study

- New aspects of estimation of the case fatality risk (CFR) have been investigated
- It has been shown that the CFR decreased over time
- The changing age distribution of the Covid-19 cases only partly explains the decrease
- The CFR in the western European countries are comparable. Remaining differences may be explained by different testing policies and case detection rates

## Transparency statement

The corresponding author affirms that this manuscript is an honest, accurate, and transparent account of the study being reported; that no important aspects of the study have been omitted; and that any discrepancies from the study as planned (and, if relevant, registered) have been explained.

## Contributorship statement

Heiko Becher had the study idea, supervised the analysis, wrote the initial draft and most parts of the final version of the manuscript

Katharina Olszewski performed the analysis, and contributed to writing of the manuscript

Sarah Wiegel supervised the analysis and contributed to writing of the manuscript

Olaf Müller contributed to the interpretation of the results and wrote parts of the manuscript

All authors approved the final manuscript

## Data sharing statement

All data used for this study are from open sources. Data are freely available under https://npgeo-corona-npgeo-de.hub.arcgis.com/datasets/d4580c810204019a7b8eb3e0b329dd6_0/data. and https://www.ecdc.europa.eu/en/publications-data/download-todays-data-geographic-distributioncovid-19-cases-worldwide.

